# Pooling saliva to increase SARS-CoV-2 testing capacity

**DOI:** 10.1101/2020.09.02.20183830

**Authors:** Anne E. Watkins, Eli P. Fenichel, Daniel M. Weinberger, Chantal B.F. Vogels, Doug E. Brackney, Arnau Casanovas-Massana, Melissa Campbell, John Fournier, Santos Bermejo, Rupak Datta, the Yale IMPACT Research Team, Charles S. Dela Cruz, Shelli F. Farhadian, Akiko Iwasaki, Albert I. Ko, Nathan D. Grubaugh, Anne L. Wyllie

**Author notes:** Correspondence (EPF); (ALW). Joint senior authors (Eli P. Fenichel, Nathan D. Grubaugh, Anne L. Wyllie).

## Abstract

Expanding testing capabilities is integral to managing the further spread of SARS-CoV-2 and developing reopening strategies, particularly in regards to identifying and isolating asymptomatic and pre-symptomatic individuals. Central to meeting testing demands are specimens that can be easily and reliably collected and laboratory capacity to rapidly ramp up to scale. We and others have demonstrated that high and consistent levels of SARS-CoV-2 RNA can be detected in saliva from COVID-19 inpatients, outpatients, and asymptomatic individuals. As saliva collection is non-invasive, extending this strategy to test pooled saliva samples from multiple individuals could thus provide a simple method to expand testing capacity.

However, hesitation towards pooled sample testing arises due to the dilution of positive samples, potentially shifting weakly positive samples below the detection limit for SARS-CoV-2 and thereby decreasing the sensitivity. Here, we investigated the potential of pooling saliva samples by 5, 10, and 20 samples prior to RNA extraction and RT-qPCR detection of SARS-CoV-2. Based on samples tested, we conservatively estimated a reduction of 7.41%, 11.11%, and 14.81% sensitivity, for each of the pool sizes, respectively. Using these estimates we modeled anticipated changes in RT-qPCR cycle threshold to show the practical impact of pooling on results of SARS-CoV-2 testing. In tested populations with greater than 3% prevalence, testing samples in pools of 5 requires the least overall number of tests. Below 1% however, pools of 10 or 20 are more beneficial and likely more supportive of ongoing surveillance strategies.

## Background

Throughout the COVID-19 pandemic, limited testing capacity in the United States has hindered both access to testing and the return of actionable test results. To control the continuing outbreaks, testing capacity must be sustainably increased and maintained for the foreseeable future. In addition, the focus of testing must shift from diagnostics to screening and continued surveillance. For this, testing capacity must not only be expanded but then also maintained to allow schools and workplaces to safely reopen - and remain open. Pooling of samples for SARS-CoV-2 testing is one resource-saving approach to increase testing capacity. This approach of “batched” testing allows multiple individuals to be tested at once to detect the presence of SARS-CoV-2 in any one of the samples^1^. Under these conditions, a negative result represents a negative test for all included samples while a positive test will initiate individual re-testing of all samples. While pooling has traditionally focused on PCR testing of extracted nucleic acid^1-3^, pooling to aid testing demands can also occur at the initial RNA extraction step, sending samples through the entire process together, right from the start. While studies have evaluated the resource-saving benefits of these approaches, the inclusion of laboratory data to help inform their approaches is rare.

We have recently demonstrated the potential of saliva as an alternative samples type for the detection of SARS-CoV-2^4,5^. Saliva can be reliably self-collected, in simple collection tubes, without the need of expensive stabilizing buffers or cold chain transport^6^. With saliva providing a mechanism to meet the needs of growing testing demands, we explored the potential of pooling saliva samples to increase sample throughput, by testing samples in pool sizes of 5, 10, and 20. Our data indicate that while pooling of saliva does decrease virus detection, it still detects the majority of infections. Pooled testing strategies do not have to be fixed, however. By modelling the overall number of tests required at different levels of virus prevalence within a population, adaptive strategies should be considered to maximize the costs-savings benefits which in turn can permit continued surveillance against virus resurgence as communities reopen.

## Results

To develop saliva pooling approaches to help meet mass testing demands, we evaluated the effects of pool sizes of 5, 10, and 20 on SARS-CoV-2 detection. We created saliva pools using samples collected from COVID-19 inpatients and healthcare workers^5^, and combined virus-positive saliva (<38 PCR cycle threshold [Ct])^7^, across a range of Ct values, with negative saliva prior to RNA extraction and RT-qPCR detection of SARS-CoV-2 RNA. All samples had previously been tested, unpooled, by RNA extraction and RT-qPCR. As expected, we found that as the pool size increased, the sensitivity decreased (pool of 5, +2.2 Ct, 95% CI 1.4, 3.0; pool of 10, +3.1 Ct, 95% CI: 2.3, 4.0; pool of 20, +3.6 Ct, 95% CI: 2.7, 4.4; **Figure 1**). Importantly, this effect on the sensitivity of detection was independent of the Ct value of the undiluted sample (Pearson’s, r=-0.004; 95% CI: −0.240, 0.233), i.e. the sensitivity loss in a sample with a higher Ct value (lower viral load) was not more than that of a sample with a lower Ct value (lower viral load). Our findings are consistent with what others have previously reported for the pooled testing of swabs^8^. However, by increasing the extraction volume (from 300 μL to 400 μL) while keeping elution volume constant, viral detection in pooled samples improved to near undiluted levels (pool of 5, −0.1 Ct, 95% CI −1.2, 1.1; pool of 10, 0.3 Ct, 95% CI −0.8, 1.5; pool of 20, 1.1 Ct, 95% CI −0.1, 2.2) (**Supplemental Figure 1**).

**Figure 1.**
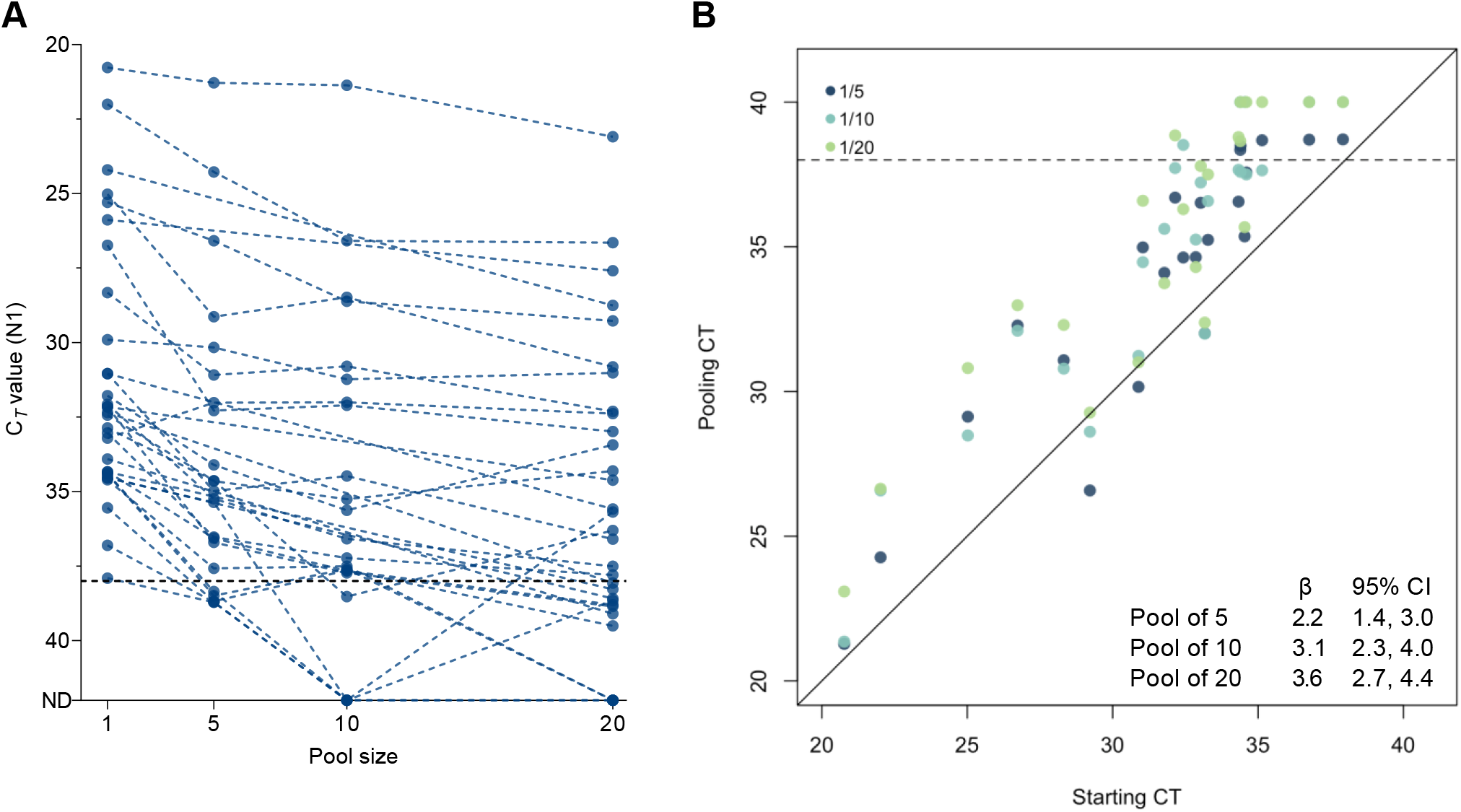
The effect of pooling on SARS-CoV-2 detection varies not only by pool size but also between samples tested. Each positive saliva sample was combined with additional negative saliva samples to form total pool sizes of 5, 10, or 20. RNA extracted from the pool was tested in RT-qPCR for SARS-CoV-2 targeting the nucleocapsid (N1). (A) As the pool size increased, as did the Ct value (dotted lines connect pools comprised of the same positive sample). Ct threshold for positivity is set to 38. Samples falling on the x-axis indicated samples from which signal was not detected (ND) in RT-qPCR. (B) We equated this change using linear regression (pool of 5, dark blue, +2.2 Ct, 95% CI 1.4, 3.0; pool of 10, light blue, +3.1 Ct, 95% CI: 2.3, 4.0; pool of 20, green, +3.6, 95% CI: 2.7, 4.4).

We also evaluated the effect of pooling post-RNA extraction and pooled RNA templates extracted from undiluted saliva samples by 5 and by 10. While we observed a similar decrease in sensitivity (pool of 5, +2.2 Ct, 95% CI: 1.7, 2.6; pool of 10, +3.1 Ct, 95% CI: 2.6, 3.6) as to when pooled prior to RNA extraction, the degree to which each sample varied was less with less overall variation as compared to pre-extraction pooling (F test, pools of 5, *p* = 0.061; pools of 10, *p* = 0.009, **Supplemental Figure 2**).

To evaluate how pooling of samples may affect the classification of an individual sample within that pool as positive or negative, we applied the regression coefficients (Ct increase) for the 5, 10, and 20 pool sizes to the Ct values from all positive saliva samples detected during our Yale IMPACT Research Studies^5^ (n = 135). We estimate that the pool sizes of 5, 10, and 20 would lead to detection sensitivities of 92.59% (95% CI: 88.89, 95.56), 88.89% (95% CI: 80.00, 91.85), and 85.19% (95% CI: 75.56, 91.11) of samples relative to that of unpooled samples (**Figure 2**).

**Figure 2.**
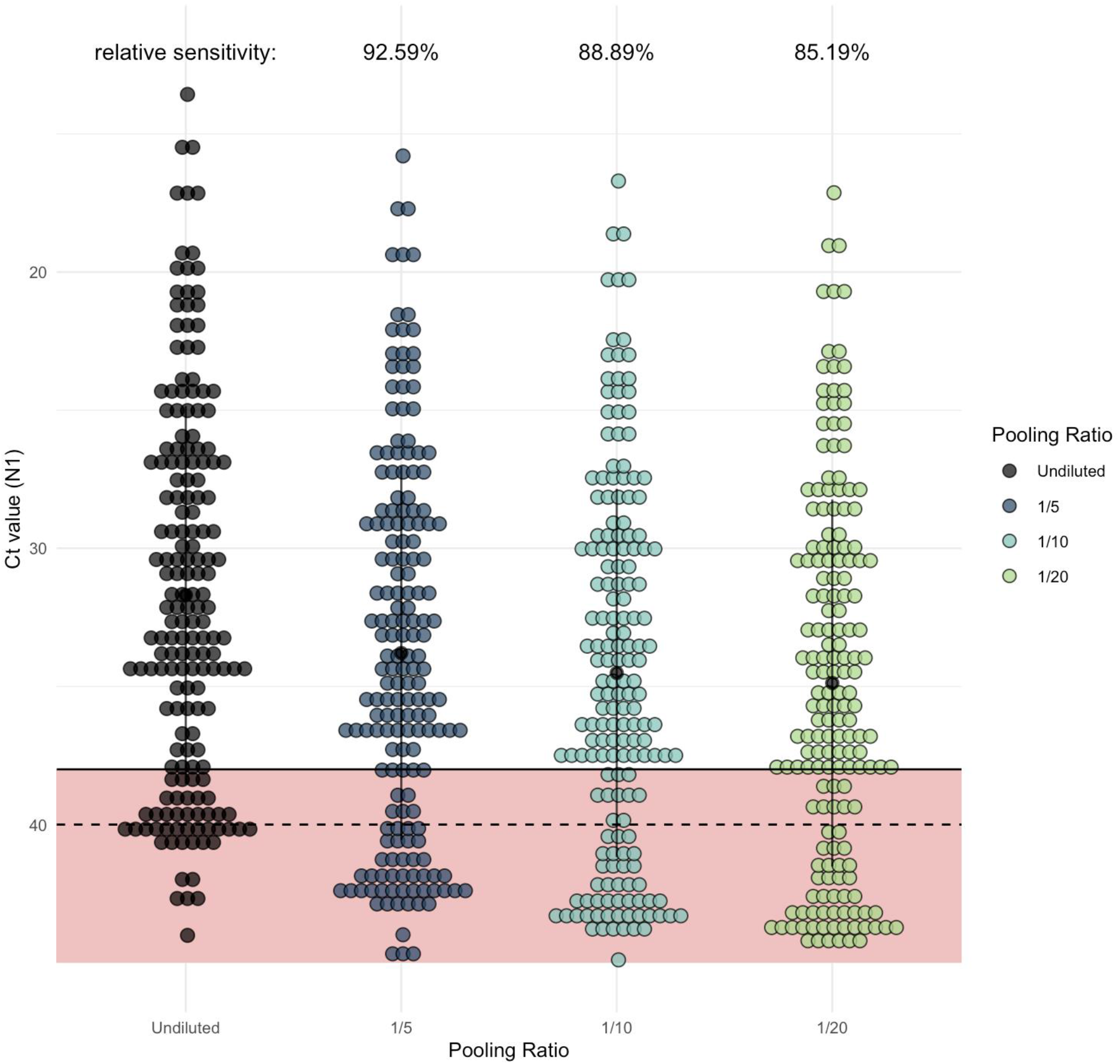
As pool size increases, more samples would be classified as negative in comparison to samples tested individually (unpooled). Each dot represents one of the Yale IMPACT saliva samples which generated signal when tested by RT-qPCR for SARS-CoV-2 N1. Of these, 135 fell below the cycle threshold (Ct) of 38 and were classified as positive for virus. The regression coefficient (representing expected Ct increase) for each of the pool sizes was added to the Ct value generated from the undiluted sample (shown in black) to determine the relative level of sensitivity for each pool size.

The goal of broad based testing is to identify people who are infectious and isolate them from the population. A cost effective strategy identifies the maximum number of infected individuals with the fewest tests. Therefore, based on the calculated relative sensitivity loss resulting from pooling, as compared to testing samples individually (**Figure 2**), we modelled the number of tests required to test a population of 10,000 (results of which scale with larger populations) with increasing prevalence of SARS-CoV-2 in pools of 5, 10, or 20 (with individual re-testing of all samples within a pool testing positive; **Figure 3A**). With these sensitivity estimates (relative to individual testing), we estimate that pooling of samples permits fewer total tests than when testing samples individually, up to a prevalence of 30%. It is possible that pools of 2 to 4 could continue to provide cost savings beyond 30%, but we did not investigate these pool sizes. Once prevalence exceeds 3% however, our analyses suggest that pooling samples by 5 results in the fewest tests required. Pools of larger sample size are more likely to test positive more often, requiring a greater number of individual samples to be retested, with more overall tests required. At a prevalence of <0.8%, we found that pools of 20 greatly reduce the number of tests needed and the cost of testing, which are important factors to ensure continued surveillance and early identification of virus resurgence in a population. If tests have a constant cost, then the cost savings for testing the population are the test cost times the change in the number of tests.

**Figure 3.**
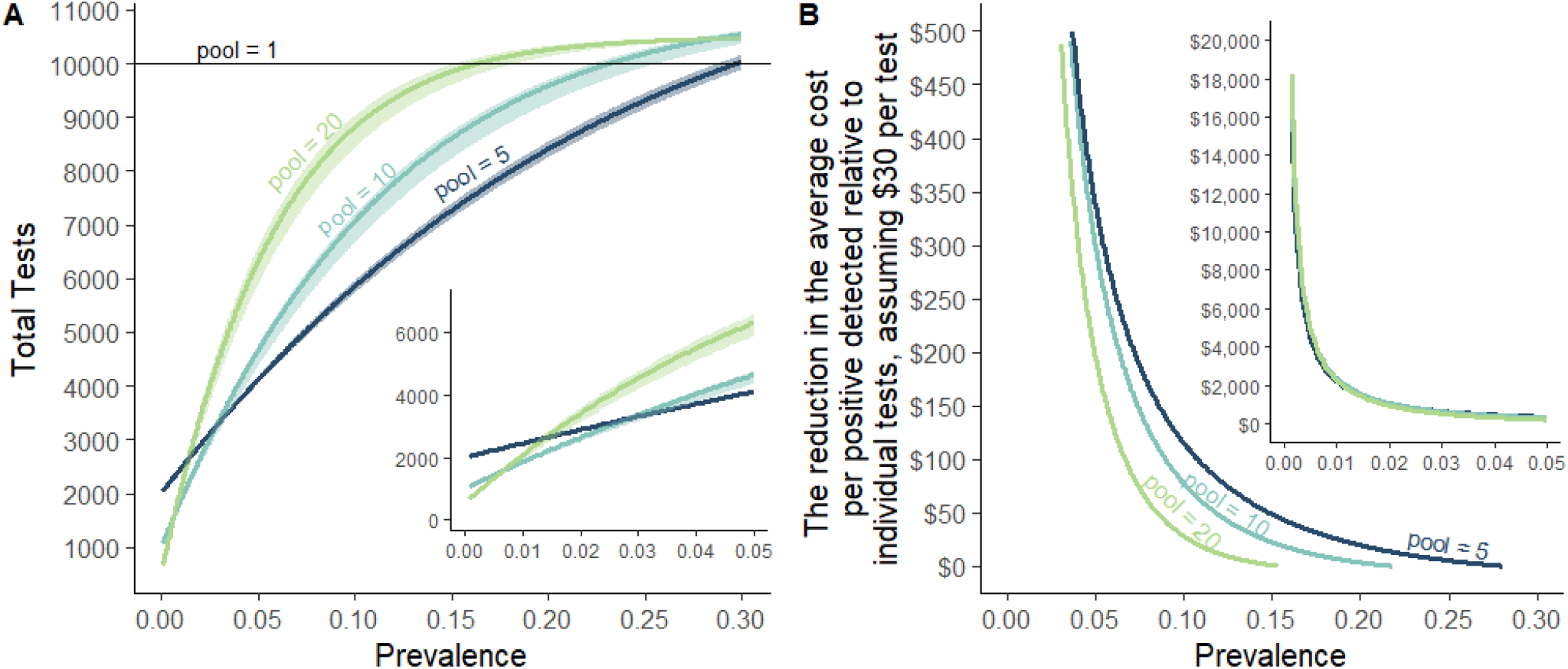
The resource-saving benefit of sample pooling for SARS-CoV-2 testing depends upon the size of the pool and the expected prevalence of SARS-CoV-2 within the population. We modeled (A) the number of tests required to test a population of 10,000 people (results qualitatively scale with population), when pooling samples by 5, 10, or 20 (and individually retesting samples within positive pools) as compared to testing samples individually (pool = 1). As prevalence increases, as will the number of pools testing positive for SARS-CoV-2, thereby increasing the required number of confirmatory tests of individual samples. Therefore, over a prevalence of 3%, pooling samples by 5 results in less overall tests required as compared to larger pool sizes. (B) At lower prevalences, such as when outbreaks have been controlled but ongoing surveillance is required, pooling samples by 10 or 20 yields substantial cost savings for the same expected level of positive detections, after accounting for sensitivity differences. Insets zoom into the region below 5% prevalence.

The purpose of testing the population is to identify infectious individuals, and different pooling designs have different sensitivities implying a different number of positives will be detected for a given population with a given prevalence. As the prevalence of the virus decreases in the tested population, we found that the cost savings through pooled testing increase (**Figure 3B**). For example, our findings show that if prevalence were 0.5%, then a population of 10,000 people can be tested with as few as 1,318 tests, including retesting of all individual saliva samples from test-positive pools. If tests cost $30 each, this represents a savings of $260,453 relative to individual testing while still identifying 43-50 individuals infected. Critically, the cost saving could allow more frequent testing and ultimately a greater identifying more infectious individuals separating them from the population at large. Adapting pools to local prevalence holds down the incremental cost. Thus, pooled testing is likely to continue to pass a benefit-cost even as prevalence falls because pool sizes can be increased. This is essential for continued surveillance as when testing samples individually, the cost of testing can be prohibitive when positive tests are rarely found. Pooling of samples can help to overcome this.

## Discussion

While pooling of samples has been widely proposed as a way to expand testing capacity for large scale screening^9-11^, there has been limited empirical evidence on pooling performance to properly inform projections of both feasibility and cost-effectiveness. Therefore, we investigated the potential of pooling saliva for SARS-CoV-2 detection in the laboratory setting, then used our findings to inform a model exploring the benefits of pooled sample testing at different rates of virus prevalence in the population. Our model demonstrates that as local outbreaks fluctuate, varying pool sizes in response will have resource-savings benefits. Taken together, pooled testing of the non-invasive and cost-effective saliva sample type facilitates extended duration and breadth of screening test strategies.

Our results suggest that in settings when prevalence in the tested population exceeds 3%, pooling samples by 5 will provide greatest savings on resources (up to 30% prevalence). While the time- and resource-savings benefits of pooled testing will always be accompanied by a decrease in the sensitivity of detection, reducing the overall number of tests required and the associated costs, permits more frequent testing, while improving overall testing capacity^14^. In turn, this will increase the capacity for testing the same individuals more often and help mitigate the loss to sensitivity^14^. Thus, the first Emergency Use Authorization by the U.S. Food and Drug Administration (FDA) for SARS-CoV-2 pooled testing (up to four swabs combined into a single test)^12^ will be the most useful in high prevalence settings.

In settings of lower prevalence (<3%), we found that larger pool sizes (10-20) will be more time and cost effective. However, the ~12-15% losses in sensitivity for pooling 10-20 samples would not likely pass the current authorization criteria by the FDA. Going forward, screening strategies need to be reviewed separately from traditional diagnostic testing, with their repeated measures taken into consideration. For strategies considering twice-weekly sampling for example (such as in the reopening plans for many U.S. colleges), even if larger pools have a lower per test sensitivity, the probability of two repeated false negative tests for any individual will often be less than the probability of a false negative from a single test from a small pool. For example, a small pool (or individual test) may have the probability of a false negative result of 2%, but only allow testing once per week. Conversely, a large pool with a per test probability of a false negative result of 14% is more likely to allow for testing twice per week. Therefore, individuals tested twice in the larger pools have a per week probability of testing falsely negative of only 1.96%. In the context of prolonged surveillance, sensitivity should be thought of as per unit time and the testing regime should be taken into account. Ultimately, the probability of a false negative should not be considered per test, but rather for a given testing regime over a specified period of time.

The results presented here are a conservative estimate for a pooled approach; the number of tests required is likely to be lower than predicted at higher prevalence, with multiple positive samples possible within a single pool. Moreover, adjustments can be made to established testing protocols to help minimize the effect of sample dilution and improve virus detection. While we replicated our extraction method by which the undiluted samples were originally tested, increasing the sample tested to the maximum input volume (400 μl) can increase the detection sensitivity. A further modification could include decreasing the RNA elution volume from 75 μl to 50 μl. In addition, while our protocol was originally set to an RT-qPCR threshold for positivity of Ct 38, we have recently expanded this to Ct 40^4^, and pooled approaches could in fact consider retesting of individual samples in pools generating any signal regardless of threshold. While we observed less variation in Ct values following pooling of RNA templates, pooling of samples prior to RNA-extraction had a similar overall change in Ct values. Due to the expense of RNA extraction, we recommend pooling prior to RNA extraction.

Pooling saliva samples for SARS-CoV-2 detection provides a mechanism to support the testing of a larger number of individuals with substantial cost savings, especially at lower levels of prevalence. Realizing these savings are important to maintain surveillance as the virus is brought under control in order to avoid resurgence - even at a very low prevalence, it is likely desirable to increase pool sizes before stopping testing altogether. Unlike standard diagnostic testing, a pooled approach for ongoing screening and surveillance is just as much about clearing non-infected individuals and providing confidence as they are about diagnosing infected individuals. Together with the ease of saliva collection, this strategy should be considered as an effective testing strategy to expand the breadth of testing and continued surveillance during the ongoing COVID-19 pandemic.

## Methods

### Sample pooling

Saliva was collected as a part of the Yale IMPACT Biorepository^5^ from COVID-19 inpatients and healthcare workers at the Yale-New Haven Hospital (Yale Human Research Protection Program Institutional Review Boards FWA00002571, Protocol ID. 2000027690)^5^. RNA was extracted and tested by RT-qPCR for SARS-CoV-2 RNA (N1)^7^.

Saliva samples were combined into pools of 5, 10, and 20. Each pool contained equal amounts of one SARS-CoV-2 positive sample (as determined by RT-qPCR) and the respective amount of SARS-CoV-2 negative samples to complete the target pool size. RNA extraction from pooled samples and RT-qPCR for SARS-CoV-2 detection were performed according to the biorepository’s standard operating procedures^5,7,15^ with either 300 μl (equating to 60 μl, 30 μl, and 15 μl of the original sample) or 400 μl (equating to 80 μl, 40 μl, and 20 μl of the original sample) total extraction input volume with RNA eluted into a total volume of 75 μl. Later, RNA extracted from saliva was tested individually or together in pool sizes of 5 or 10 and tested in RT-qPCR for SARS-CoV-2 detection^7^.

### Statistical analyses

#### Sensitivity analyses

We fit a linear regression to the experimental pooling data to model the change in Ct values of positive samples following pooling. Let ‘ΔCt’ be the change in Ct value of pooled samples and let ‘ratio’ be the categorical ratio of pool size (i.e. 1/5, 1/10, 1/20). Analyses were done separately by input volume in order to determine the effect of pool size under both 300 μl and 400 μl extraction conditions. This equation was used, separately, for both pre-extraction saliva and post-extraction RNA pooling. Ratio in this model can be interchanged with “condition” for the model of the 1/20 PBS and water dilution data.

We found that the change in Ct value post-pooling was independent of the Ct value of the undiluted sample (Pearson’s, r=-0.004; 95% CI: −0.240, 0.233), thus it was not included in the model. Confidence intervals were generated by simulating from the covariance matrix of the parameters from the fitted model using the mvrnorm function in the R package “MASS”^16^, and quantile functions.

### Modeling the resource-saving benefit of sample pooling for SARS-CoV-2 testing

If samples are independent of each other, pulled from the same population, and that anyone in a test-positive pool needs to be re-tested individually, then binomial sampling theory provides the tool to compute the number of tests needed, which has been used for over half a century^17,18^. The number of positive groups is *P =* [*1 −* (*1 − σ*(*g*)*m*)*^g^*]*N*, given a total test population of size *N* that is divided into groups of size *g*, with a population prevalence of infection of *m* and a test sensitivity *σ*(*g*), where sensitivity can be a function of group size. The total number of tests need is 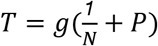. Functions to implement these calculations are available at https://github.com/efenichel/pooled-saliva-testing.

Further statistical analyses were conducted in GraphPad Prism 8.0.0 as described in the text and figure legends.

### Yale IMPACT Research Team authors

Kelly Anastasio, Michael H. Askenase, Maria Batsu, Sean Bickerton, Kristina Brower, Molly L. Bucklin, Staci Cahill, Yiyun Cao, Edward Courchaine, Giuseppe DeIuliis, Rebecca Earnest, Bertie Geng, Ryan Handoko, Christina A. Harden, Chaney C. Kalinich, William Khoury-Hanold, Daniel Kim, Lynda Knaggs, Maxine Kuang, Eriko Kudo, Melissa Linehan, Peiwen Lu, Alice Lu-Culligan, Anjelica Martin, Irene Matos, David McDonald, Maksym Minasyan, Adam J. Moore, M. Catherine Muenker, Nida Naushad, Allison Nelson, Jessica Nouws, Abeer Obaid, Camila Odio, Ji Eun Oh, Saad Omer, Isabel M. Ott, Annsea Park, Hong-Jai Park, Xiaohua Peng, Mary Petrone, Sarah Prophet, Tyler Rice, Kadi-Ann Rose, Lorenzo Sewanan, Lokesh Sharma, Denise Shepard, Mikhail Smolgovsky, Nicole Sonnert, Yvette Strong, Codruta Todeasa, Maria Tokuyama, Jordan Valdez, Sofia Velazquez, Arvind Venkataraman, Pavithra Vijayakumar, Elizabeth B. White, Yexin Yang.

## Data Availability

The data that support the findings of this study are available from the corresponding author [ALW] upon reasonable request.

## Acknowledgements

We gratefully acknowledge the study participants for their time and commitment to the study. We thank all members of the clinical team at Yale-New Haven Hospital for their dedication and work which made this study possible. We also thank S. Taylor and P. Jack for technical discussions.

## Funding

This study was funded by the Huffman Family Donor Advised Fund, Fast Grant funding support from the Emergent Ventures at the Mercatus Center, George Mason University, the Yale Institute for Global Health, Yale School of Medicine, NIAID U19 AI08992 and the Beatrice Kleinberg Neuwirth Fund. CBFV is supported by NWO Rubicon 019.181EN.004.

## Competing interests

ALW has received research funding through grants from Pfizer to Yale and has received consulting fees for participation in advisory boards for Pfizer and PPS Health. DMW has received consulting fees from Pfizer, Merck, GSK, and Affinivax and has received research funding through grants from Pfizer to Yale.

## Supplemental Figures

**Supplemental Figure 1.**
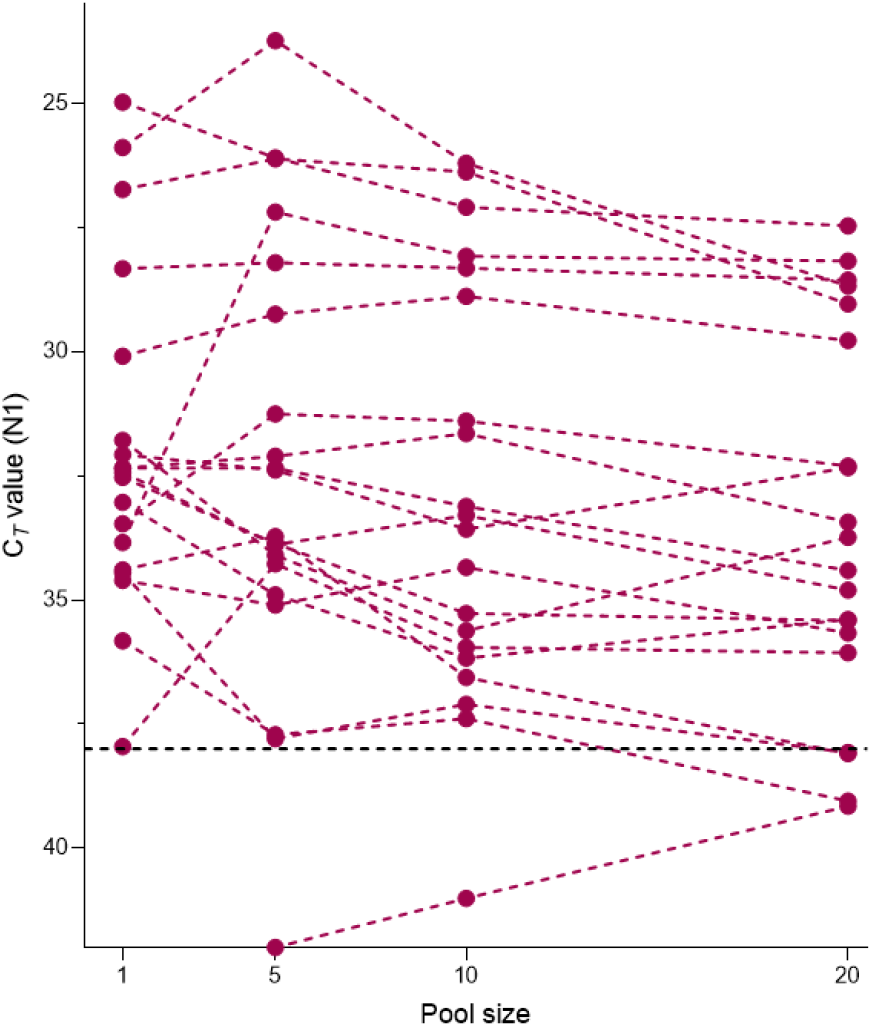
Cycle threshold (Ct) values of saliva samples tested individually (pool size = 1) at a total volume of 300 μL, or when diluted with an increasing number of negative samples (total pool sizes of 5, 10 and 20) and a total extraction volume of 400 μL. When extracting from 400 μL volumes of pooled samples, we observed improved detection (pool of 5, −0.1 Ct, 95% CI −1.2, 1.1; pool of 10, 0.3 Ct, 95% CI −0.8, 1.5; pool of 20, 1.1 Ct, 95% CI −0.1, 2.2; linear regression). Dotted lines connect pools comprised of the same positive sample. Ct threshold for positivity is set to 38. Samples falling below the x-axis indicated samples from which signal was not detected in RT-qPCR.

**Supplemental Figure 2.**
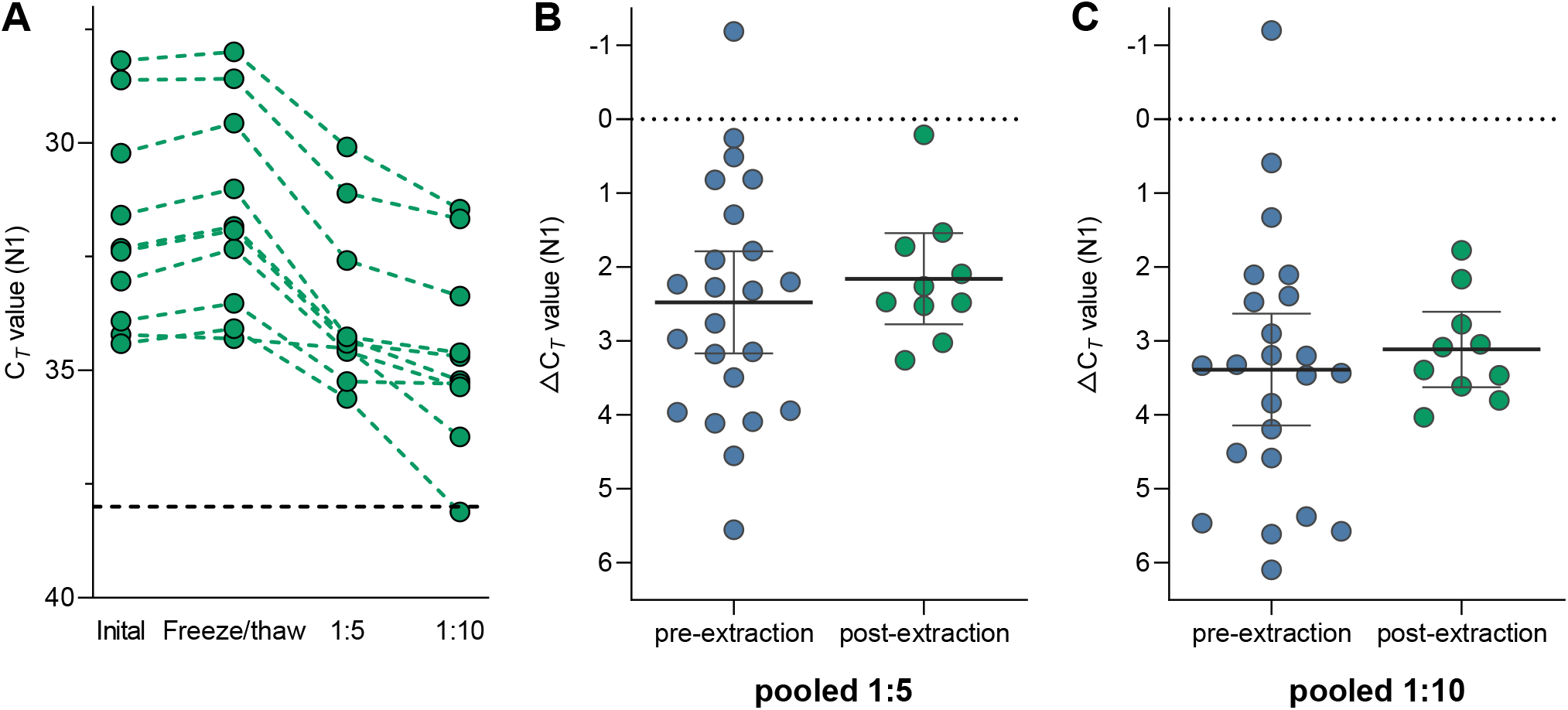
Less variation in cycle threshold (Ct) values when pooling RNA templates. (**A**) Ct values of SARS-CoV-2 positive RNA extracted from saliva samples when tested individually (pool size = 1) on day of sample collection (initial) and following storage of RNA at −80°C (freeze/thaw), or when diluted with 4 or 9 SARS-CoV-2 negative RNA templates (total pool sizes of 5 and 10). Dotted lines connect pools comprised of the same positive sample. While the median change in Ct value was comparable whether pooling samples or RNA templates by (**B**) 5 (Mann-Whitney, *p* = 0.499) or (**C**) 10 (Mann-Whitney, *p* = 0.556), pooling of samples resulted in more varied Ct changes (F test, *p* = 0.061 and *p* = 0.009, respectively).

## Notes

### Author Declarations

All study participants were enrolled and sampled in accordance to the Yale University HIC-approved protocol #2000027690. Demographics, clinical data and samples were collected after the study participant had acknowledged that they had understood the study protocol and signed the informed consent. All participant information and samples were collected in association with non-individually identifiable study identifiers. All necessary patient/participant consent has been obtained and the appropriate institutional forms have been archived.

